# Predictors of modern contraceptive use among reproductive-aged women (15-49 years) in Mozambique: Evidence from Mozambique Demographic and Health Survey

**DOI:** 10.64898/2026.09.21.26363579

**Authors:** Nahin Shakurun, Fernanda Andre, Nazeem Muhajarine

## Abstract

**Background:** Modern contraceptive use remains low in many sub-Saharan African countries, including Mozambique, limiting progress towards sexual and reproductive health and Sustainable Development Goals (SDGs). Understanding the prevalence, determinants, and inequalities in modern contraceptive use based on Mozambique-wide national survey is critical to informing effective family planning interventions.

**Method:** We analyzed the most recent data from the Mozambique Demographic and Health Survey (DHS), 2022-2023, focusing on a total of 11,783 reproductive-aged women (15-49 years). The outcome variable was current use of modern contraceptives. Several individual, household-related and community determinants of modern contraceptives were examined employing multivariable logistic regression. Adjusted odds ratio with corresponding confidence intervals and p-values were reported. Erreygers Concentration Index (ECI) and concentration curves were used to assess wealth-and empowerment-related inequalities. All analyses were conducted on weighted data.

**Result:** Overall, 27.6% of women reported current use of modern contraceptive methods. Younger age, higher education, being employed, higher parity, greater wealth, and a partner with higher education were positively associated with modern contraceptive use. Significant wealth-related (ECI = 0.32, p < 0.001) and women’s empowerment-related (ECI = 0.26, p < 0.001) inequalities were observed, indicating that wealthier and more empowered women had higher use of modern contraceptive methods.

**Conclusion:** Modern contraceptive use in Mozambique is suboptimal and influenced by a combination of individual, household, and contextual factors, with marked socioeconomic and empowerment-related disparities. Policy interventions should focus on improving access to and encouraging use of contraceptives for economically disadvantaged and women with low-empowerment, promoting women’s autonomy, enhancing male involvement, and leveraging media and community-based programs to increase awareness and uptake. Equity-focused strategies are essential to achieving national and global family planning targets.

## INTRODUCTION

The World Health Organization (WHO) has proposed that the prevalence of contraceptive use as one of the key national and global indicators of reproductive health.(1) There is a wide range of contraceptive methods available today. These include permanent methods and long-acting irreversible contraceptives.(2) According to the WHO, modern contraceptive methods refer to hormonal or non-hormonal methods, including pills, intrauterine devices (IUDs), implants, injections, condoms, and sterilization.(1,3) In the last three decades, the use of contraceptives has greatly increased worldwide.(4,5) However, access to and utilization of contraceptives remain a major developmental challenge in most African countries, especially among women of reproductive age (15-49 years).(6) Contraceptive use is not only a significant individual component of reproductive health, but also a crucial factor for reducing fertility, spacing of births, particularly in limited-resource settings and escaping from the poverty-fertility burden.

Moreover, contraceptive use empowers women to take control of their reproductive health. It reduces the risk of unwanted pregnancies and minimizes psychological, cultural, social, financial, and health outcomes burdens. Modern contraceptives also play a vital role in reducing maternal and child morbidity and mortality rates through preventing high-risk pregnancies and promoting proper birth spacing.(3,7,8) Furthermore, global estimates indicate that 218 million women in low-and middle-income countries (LMICs) have unmet need for modern contraceptives. This greatly contributes to poor maternal outcomes, including high rates of unwanted pregnancies, unsafe abortions, and maternal deaths.(9)

In sub-Saharan Africa, only 29.6% of women of reproductive age who wish to avoid pregnancy have access to and utilize modern contraceptives, contrasting the global average of 65%.(10,11) Over time, Mozambique has made significant strides in access to and utilization of modern contraceptives. The prevalence rates among reproductive-age women rose from 13 per cent in 2003 to 20 per cent in 2015, and to 26 per cent in 2022.(13,14) This upward trend highlights the global focus on modern contraceptives as essential for reproductive healthcare services, well-being, and socioeconomic development.(3,13,14)

Use of modern contraception is a key identifiable indicator to achieving Sustainable Development Goals (SDGs).(15) Global commitments such as the SDGs include specific targets for advancing reproductive health through access to and use of modern contraceptives. The SDGs indicator 3.7.1 on contraceptive use calls on countries to ensure universal access to sexual and reproductive health-care services. This includes family planning, sexual health education, and integrating reproductive health into national health strategies and programmes.(16)

Previous research conducted elsewhere has suggested that misconceptions, cultural competency and religious myths greatly influence modern contraception use.(17,18) Additional studies have highlighted that intimate partner-related factors, sociodemographic characteristics, parity, media exposure, fear of side effects, and health literacy are associated with modern contraceptive use. (10,18,19) Although a number of studies have recognized the importance of modern methods of contraception, few directly examined the factors associated with its use among Mozambican women of reproductive age. This study addresses that gap by estimating the prevalence of modern contraceptive use and identifying its determinants among women aged 15-49 years in Mozambique.

## METHOD

### Study design

The study utilized data from the Mozambique Demographic and Health Survey (DHS) 2022-23, a nationwide population-based survey, including all ten provinces (Niassa, Cabo Delgado, Nampula, Zambezia, Tete, Manica, Sofala, Inhambane, Gaza, and Maputo), and the capital city of Maputo, which has provincial status.(20) A two-stage stratified sampling design was followed for data collection from the households. In the first stage, clusters (enumeration areas or EAs) were selected, based on the 2017 IV General Census of Population and Housing (IV RGPH). A total of 619 EAs were chosen with probability-proportional-to-size, measured by the number of households in each explicit stratum. In the second stage, 26 households were systematically selected with equal selection probabilities from each EA. Based on this procedure, 16,045 households, representative of all households in each EA, were selected for data collection. All women aged 15-49 years who were residents or were visitors in the household the night before the interviews were eligible for the interview. For this analysis we have included reproductive age women (15-49-years) from individual record (IR) data file.(21) Women who were pregnant at the time of interview or were declared not able to have children were excluded from this analysis. Details of the participants selection for this analysis are presented in supplementary file (**Figure S1)**.

### Variables

#### Outcome variable

The outcome variable for our study was current use of modern contraceptives by the reproductive-aged women in Mozambique (at the time of the survey). Based on the definition of Hubacher and Trussell,(22) we considered modern contraceptive methods to include: oral contraceptive pills, intrauterine devices (IUD), injectables, diaphragm, male/female condoms, female/male sterilization, implants/Norplant, foam or jelly, and emergency contraception. The outcome variable was dichotomized as “use of a modern contraceptives (coded as “1”) and non-use of modern contraceptives (coded as “0”).

#### Individual factors

These included demographic and socio-economic variables such as women’s age (15–19, 20–24, 25–29, 30–34, 35–39, 40–44, 45–49) women’s educational level (No formal education, primary school, secondary school, higher), marital/partnered status (Never in union, partnered, non partnered), employment status (Employed, Unemployed), religion (Catholic, Islam, Zion, Evangelical/Pentecostal, and Others), women’s age at first sex (<15 years, 15-19 years, 20 years and above), number of living children (no children, 1-2 children, 3-4 children, five or more children), husband/partner’s education level (No formal education, primary school, secondary school, higher). Other variables were decision-maker for the contraceptive use (Respondent, Husband/partner, Joint decision, Other) and heard about family planning on media in the past 12 months, which, in turn, was derived from a combination of three variables; a) heard about family planning on radio, b) on television or c) from reading newspaper/magazine. The combined variable was dichotomized into No (0= not from any) and Yes (1= at least from one source) categories. The community characteristics include the type of place of residence (Urban, Rural) and region of residence (11 administrative regions of Mozambique). Details of the categorization of the individual level variables are presented in supplementary file **(Table S1)**.

#### Social equity marker

To investigate the social inequalities in utilization of modern contraceptives we have focused on two key indicators: household wealth status, and women’s empowerment index. (i) Household wealth index: This index serves as a proxy for economic status and is derived through principal component analysis (PCA). It incorporates various household characteristics such as asset ownership (e.g., televisions, bicycles, automobiles), construction materials of the dwelling, access to water and sanitation, and types of fuel used for cooking. The resulting continuous wealth score is segmented into five categories: Poorest (Quintile 1), Poorer (Quintile 2), Middle (Quintile 3), Richer (Quintile 4), and Richest (Quintile 5). This variable is precomputed and readily available within the dataset. (ii) Women’s empowerment index: For this study, we have adapted the Survey-based Women’s Empowerment Index (SWPER) to better align with our dataset and research objectives.(23) Due to overlaps between the original SWPER components and our selected variables, we constructed a tailored version using 11 indicators that span three dimensions of empowerment: autonomy in decision making, social independence, and attitude towards gender-based violence. Rather than generating separate scores for each domain, we applied PCA to create a unified empowerment score. This composite measure was then divided into five quintiles, mirroring the categorization used for the wealth index. Details of this variable and its categorization are presented in supplementary file (**Table S2)**.

### Statistical analysis

This study followed Guide to DHS statistics-8 (24) to perform data coding, cleaning and analysis. Frequencies and weighted percentages were employed to describe the study variables. Pearson’s Chi-square tests were applied to investigate the association between modern contraceptive use and each independent variable, estimating the unadjusted odds ratios (OR) with its corresponding 95% confidence intervals (CIs) (supplementary **table S4**). All variables with a bivariate association at p<0.20 were included in the multivariable logistic regression model. Finally, the Hosmer-Lemeshow test was used to determine the model’s goodness-of-fit. All adjusted odds ratios, their 95% CIs, and corresponding p-values are reported.

Multicollinearity between independent variables was checked using variance inflation factor (VIF). None of the variables showed multicollinearity except marital status which was removed from the multivariable model. To assess household wealth and women’s empowerment-related disparities, in modern contraceptive use, we employed the Concentration Index (CI) and Concentration Curve (CC) approaches. These tools are widely recognized to assess health inequalities. The CI produces values ranging from −1 to +1: A positive CI suggests that access is skewed toward wealthier individuals, with the CC appearing below the 45-degree line of equality. A negative CI reflects access favoring poorer groups, where the CC lies above the reference line. A CI of zero denotes perfect equity, with the CC aligning directly along the 45-degree diagonal.(25) Since the outcome variable was binary in nature, Erreygers’s correction method was followed to bound the Concentration Index value between +1 to -1.(26)

All the analyses incorporated an adjustment for sampling design using sampling weights, clustering and stratification. A complete case analysis was conducted in which only variables with complete values were used in the model. The data analysis was conducted using Stata version 18.5 (StataCorp, College Station, TX, USA).

### Ethical considerations

The DHS surveys have been reviewed and approved by Inner City Fund (ICF) Institutional Review Board (IRB) as well as Ethics Boards of partner organizations of the various countries such as the Ministries of Health. The DHS follows the standards for ensuring the protection of respondents’ privacy. ICF International ensures that the survey complies with the U.S. Department of Health and Human Services’ regulations for the protection of human subjects. This was a secondary analysis of data; therefore, no further approval was required since the data is available in the public domain. Further information about the DHS data usage and ethical standards are available at https://dhsprogram.com/methodology/Protecting-the-Privacy-of-DHS-Survey-Respondents.cfm

## RESULTS

### 3.1 Sample characteristics

**Table 1** shows the sociodemographic characteristics of women aged 15-49 years. The majority were aged 15-24 years (44.0%) and resided in rural areas (60.1%). Most had primary education (41.9%), were not working (69.3%), and were partnered (62.6%). Catholic (29.8%) and Evangelical/Pentecostal (28.8%) were the predominant religions. Over half (56.9%) reported sexual debut before age 15, and 33.0% had 1-2 living children. Women in the lowest empowerment quintile comprised 23.4%, while 25.6% belonged to the richest wealth quintile. Regarding husbands’ education, 36.03% had no formal schooling. Media exposure to family planning was reported by 41.2%. Contraceptive decisions were mostly joint (34.5%). Regional distribution was highest in Nampula (23.0%) and Zambezia (16.8%), with lower representation from southern provinces.

**Table 1:**
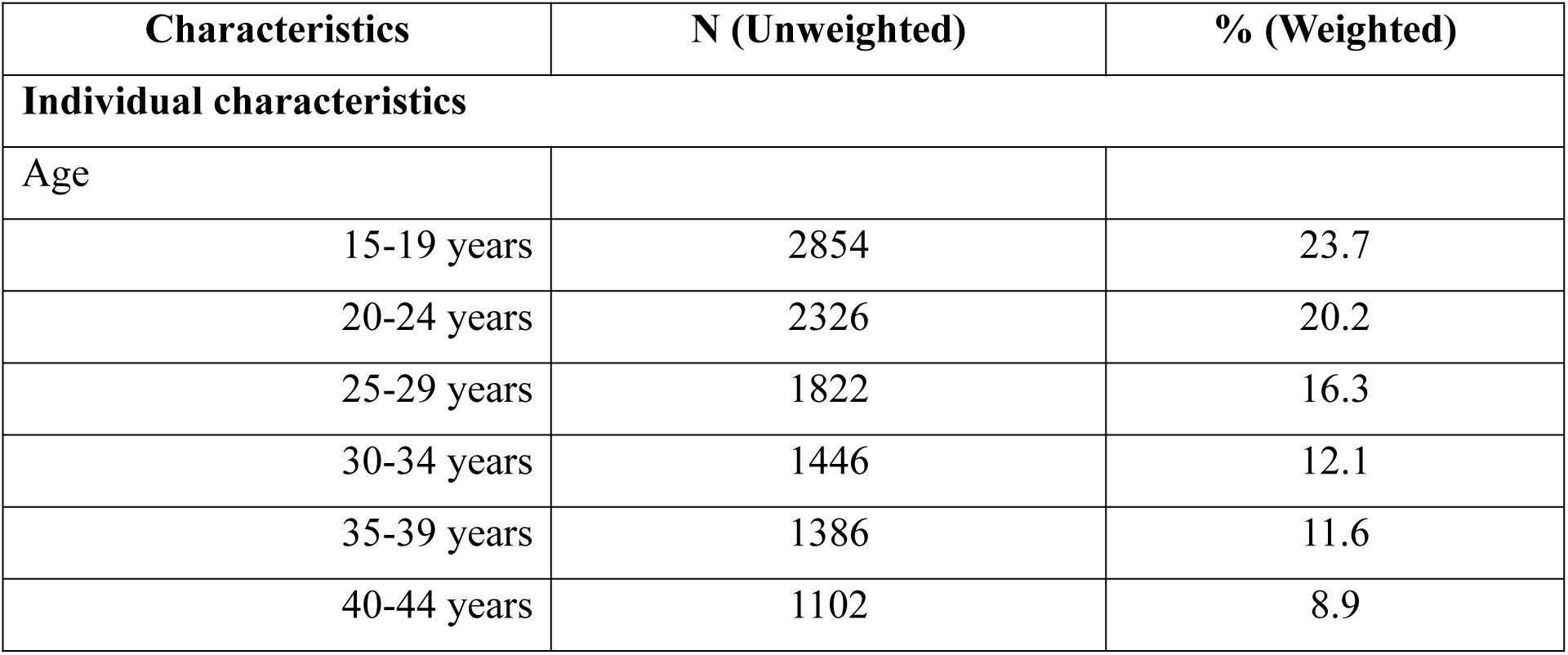
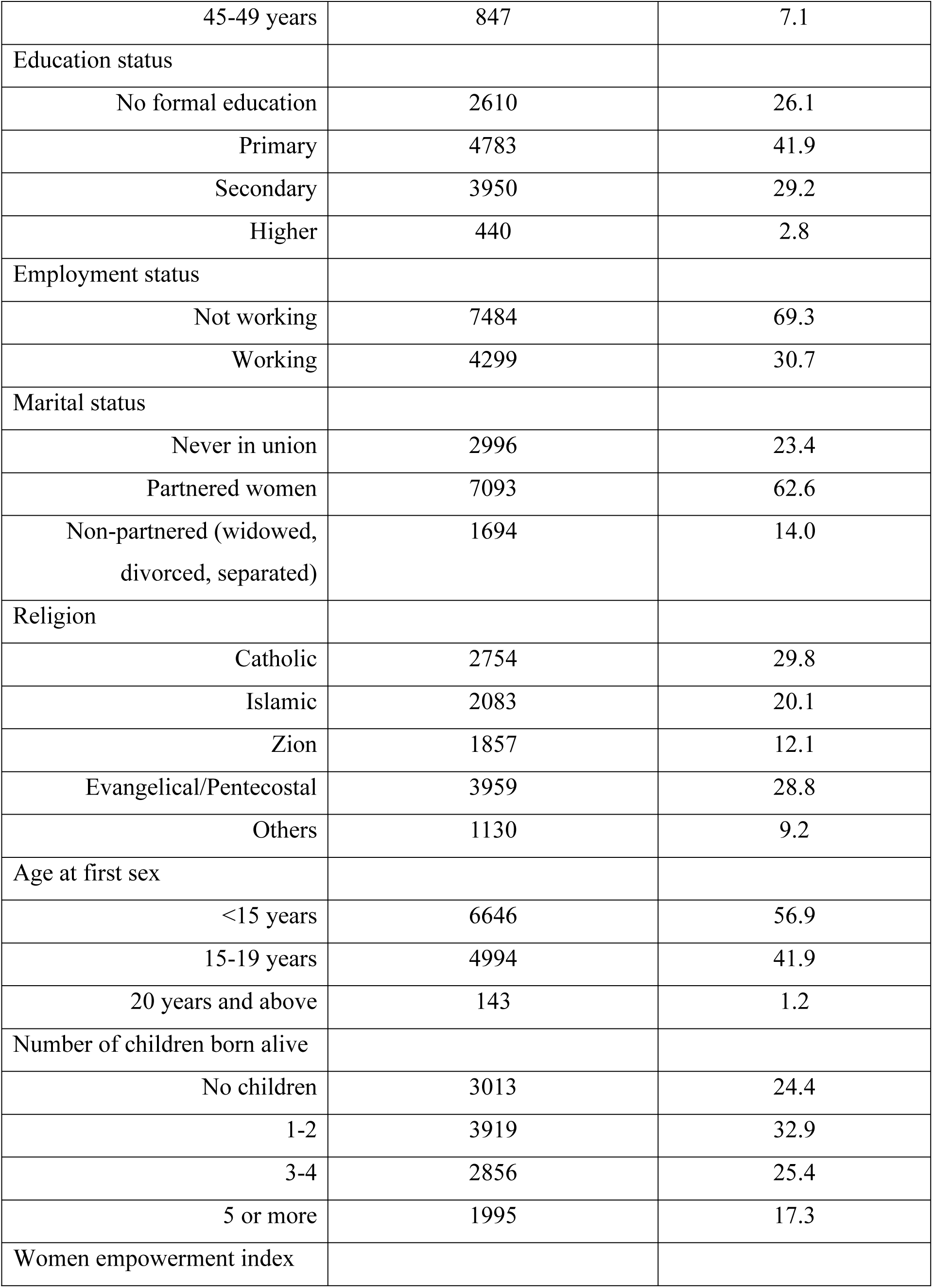
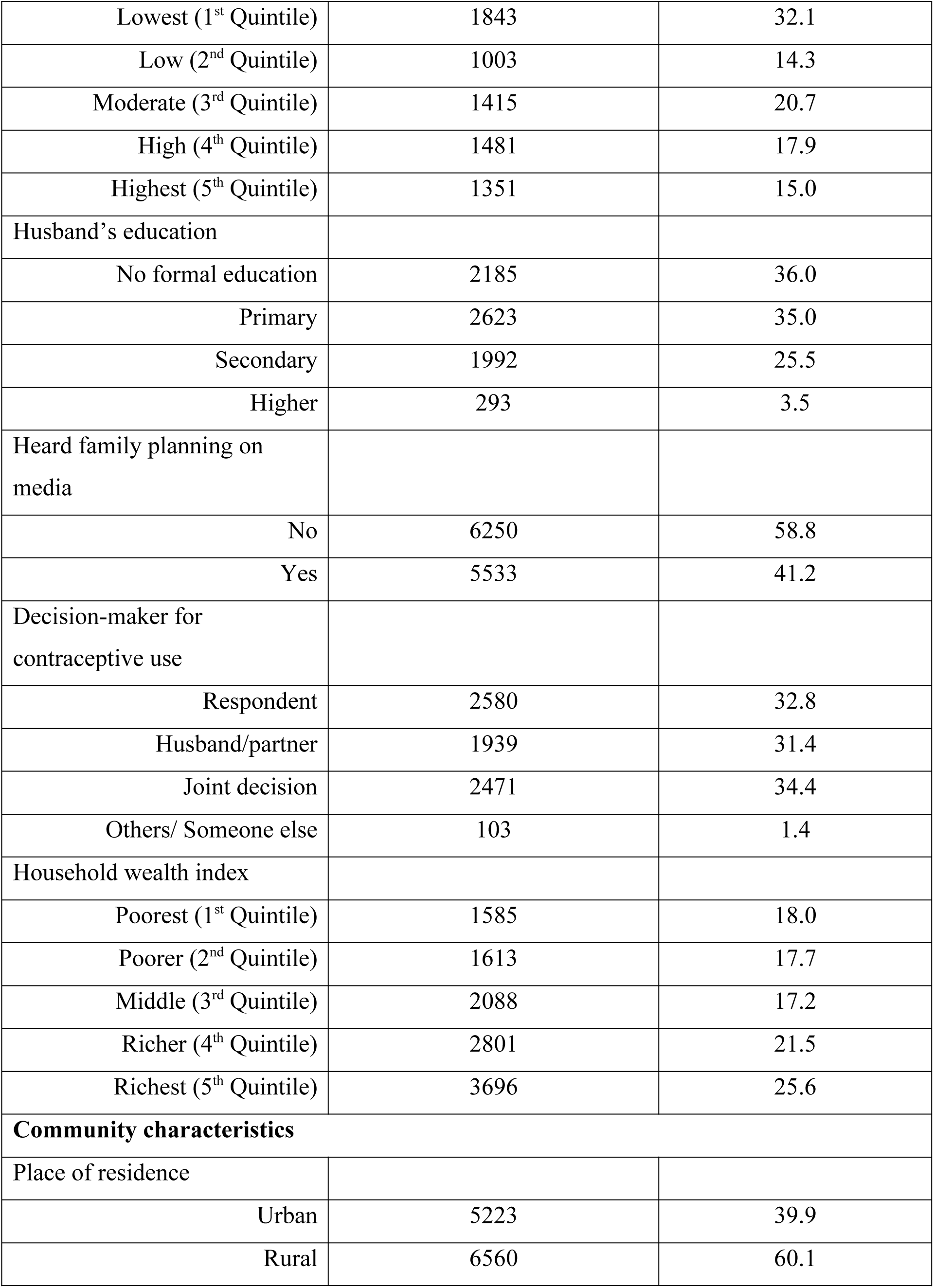
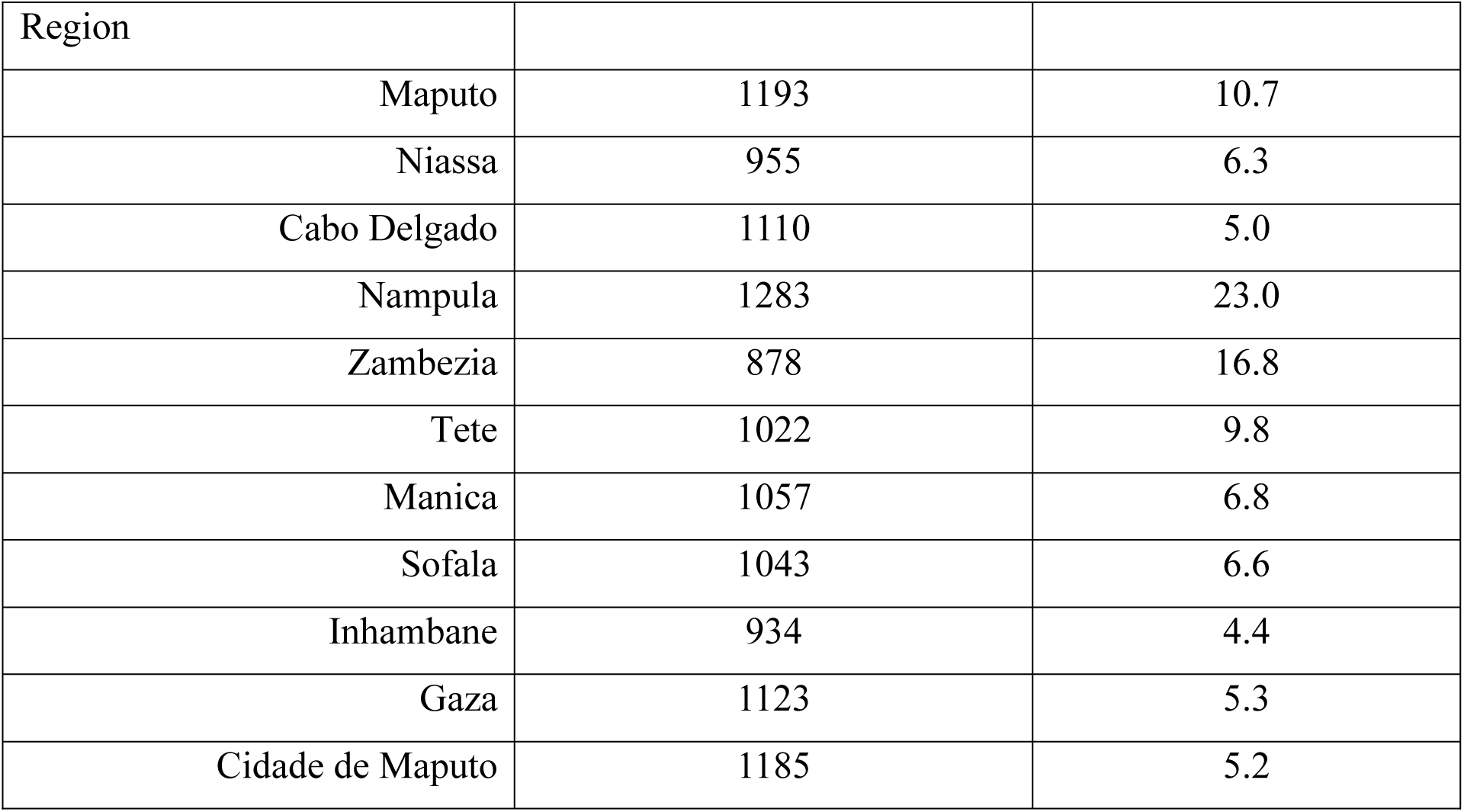
Background characteristics of the study participants: Mozambique DHS, 2022-23.

| Characteristics | N (Unweighted) | % (Weighted) |
| --- | --- | --- |
| **Individual characteristics** |  |  |
| Age |  |  |
| 15-19 years | 2854 | 23.7 |
| 20-24 years | 2326 | 20.2 |
| 25-29 years | 1822 | 16.3 |
| 30-34 years | 1446 | 12.1 |
| 35-39 years | 1386 | 11.6 |
| 40-44 years | 1102 | 8.9 |
| 45-49 years | 847 | 7.1 |
| Education status |  |  |
| No formal education | 2610 | 26.1 |
| Primary | 4783 | 41.9 |
| Secondary | 3950 | 29.2 |
| Higher | 440 | 2.8 |
| Employment status |  |  |
| Not working | 7484 | 69.3 |
| Working | 4299 | 30.7 |
| Marital status |  |  |
| Never in union | 2996 | 23.4 |
| Partnered women | 7093 | 62.6 |
| Non-partnered (widowed,<br>divorced, separated) | 1694 | 14.0 |
| Religion |  |  |
| Catholic | 2754 | 29.8 |
| Islamic | 2083 | 20.1 |
| Zion | 1857 | 12.1 |
| Evangelical/Pentecostal | 3959 | 28.8 |
| Others | 1130 | 9.2 |
| Age at first sex |  |  |
| <15 years | 6646 | 56.9 |
| 15-19 years | 4994 | 41.9 |
| 20 years and above | 143 | 1.2 |
| Number of children born alive |  |  |
| No children | 3013 | 24.4 |
| 1-2 | 3919 | 32.9 |
| 3-4 | 2856 | 25.4 |
| 5 or more | 1995 | 17.3 |
| Women empowerment index |  |  |
| Lowest (1<sup>st</sup> Quintile) | 1843 | 32.1 |
| Low (2<sup>nd</sup> Quintile) | 1003 | 14.3 |
| Moderate (3<sup>rd</sup> Quintile) | 1415 | 20.7 |
| High (4<sup>th</sup> Quintile) | 1481 | 17.9 |
| Highest (5<sup>th</sup> Quintile) | 1351 | 15.0 |
| Husband's education |  |  |
| No formal education | 2185 | 36.0 |
| Primary | 2623 | 35.0 |
| Secondary | 1992 | 25.5 |
| Higher | 293 | 3.5 |
| Heard family planning on media |  |  |
| No | 6250 | 58.8 |
| Yes | 5533 | 41.2 |
| Decision-maker for contraceptive use |  |  |
| Respondent | 2580 | 32.8 |
| Husband/partner | 1939 | 31.4 |
| Joint decision | 2471 | 34.4 |
| Others/ Someone else | 103 | 1.4 |
| Household wealth index |  |  |
| Poorest (1<sup>st</sup> Quintile) | 1585 | 18.0 |
| Poorer (2<sup>nd</sup> Quintile) | 1613 | 17.7 |
| Middle (3<sup>rd</sup> Quintile) | 2088 | 17.2 |
| Richer (4<sup>th</sup> Quintile) | 2801 | 21.5 |
| Richest (5<sup>th</sup> Quintile) | 3696 | 25.6 |
| **Community characteristics** |  |  |
| Place of residence |  |  |
| Urban | 5223 | 39.9 |
| Rural | 6560 | 60.1 |

| Region |  |  |
| --- | --- | --- |
| Maputo | 1193 | 10.7 |
| Niassa | 955 | 6.3 |
| Cabo Delgado | 1110 | 5.0 |
| Nampula | 1283 | 23.0 |
| Zambezia | 878 | 16.8 |
| Tete | 1022 | 9.8 |
| Manica | 1057 | 6.8 |
| Sofala | 1043 | 6.6 |
| Inhambane | 934 | 4.4 |
| Gaza | 1123 | 5.3 |
| Cidade de Maputo | 1185 | 5.2 |

### 3.2 Prevalence of modern contraceptive use by reproductive-aged women in Mozambique

The prevalence of modern contraceptive utilization among women of reproductive age was 27.6% (supplementary **table S3). Figure 1** illustrates the geographic distribution of varying prevalence of modern contraceptive use among women in Mozambique. The highest prevalence was found in Maputo (55.0%), and the lowest was found in Zambezia (12.2%).

**Figure 1:**
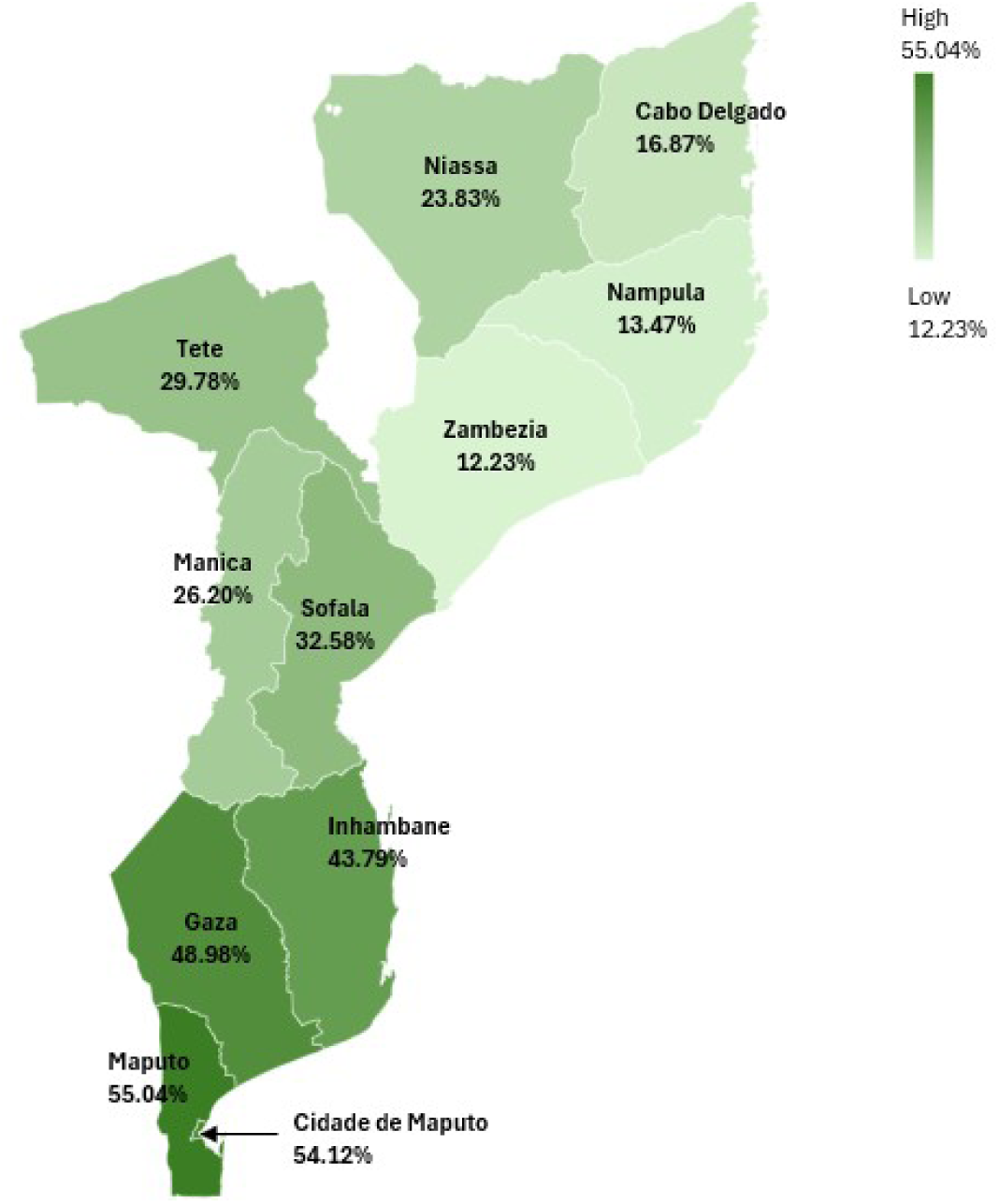
Geographical distribution of modern contraceptive use by reproductive-aged women in Mozambique (presented as weighted percentages)

### 3.3 Factors associated with utilization of modern contraceptives in reproductive aged women in Mozambique

**Table 2** shows adjusted odds ratios (aORs) from the multivariable logistic regression model examining individual, household, and community-related factors associated with modern contraceptive use among reproductive-aged women in Mozambique.

**Table 2:**
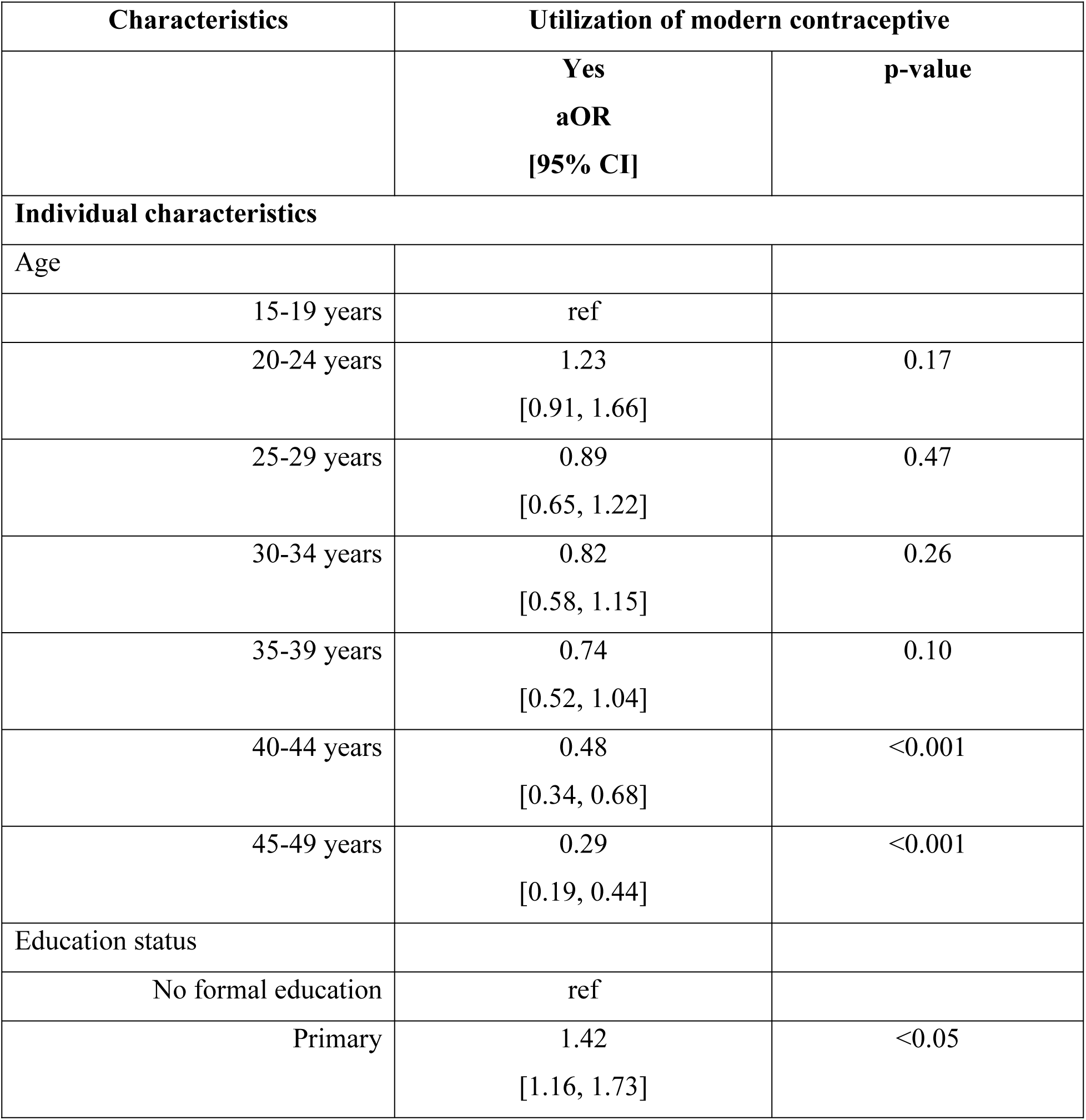
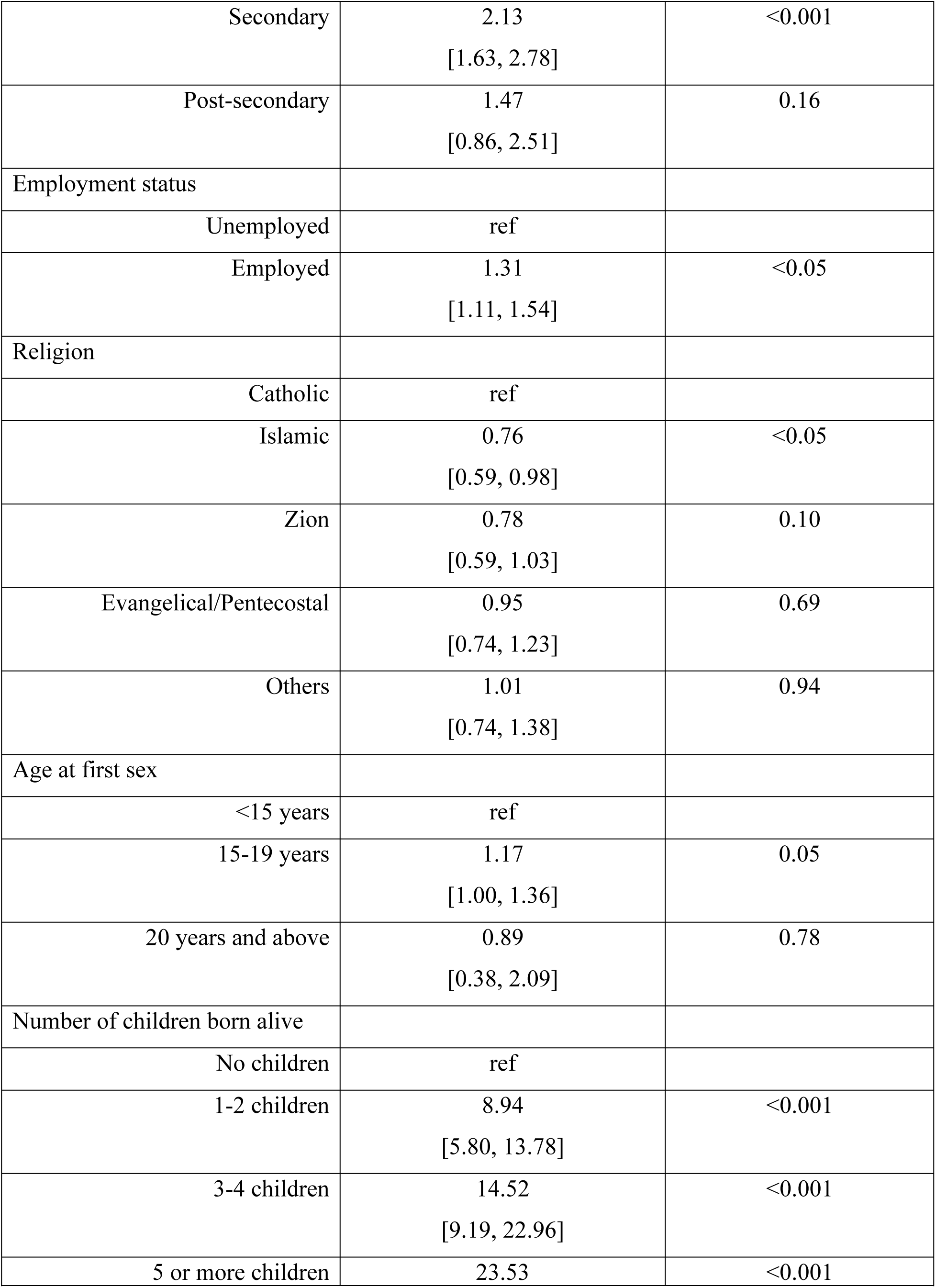
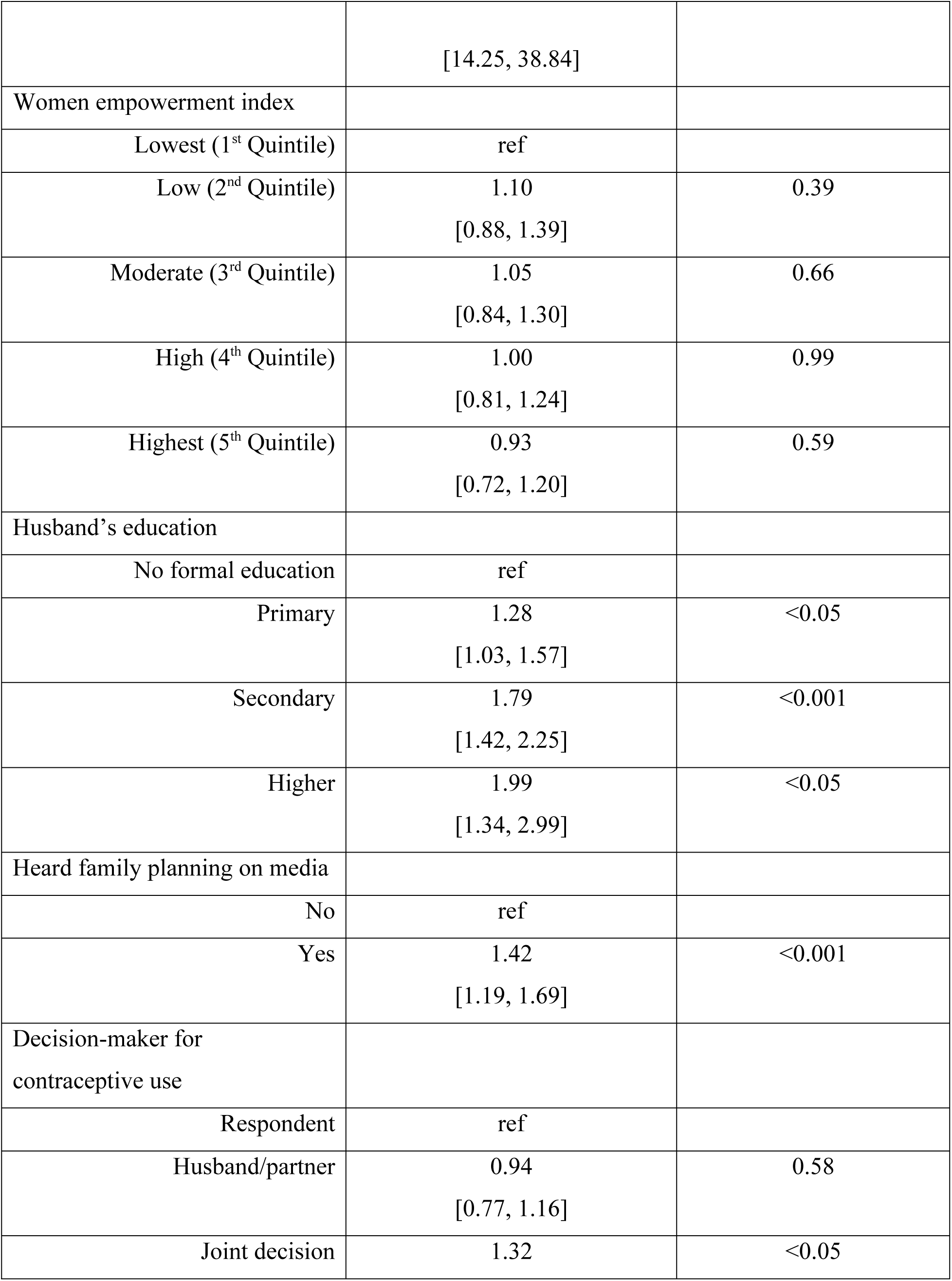
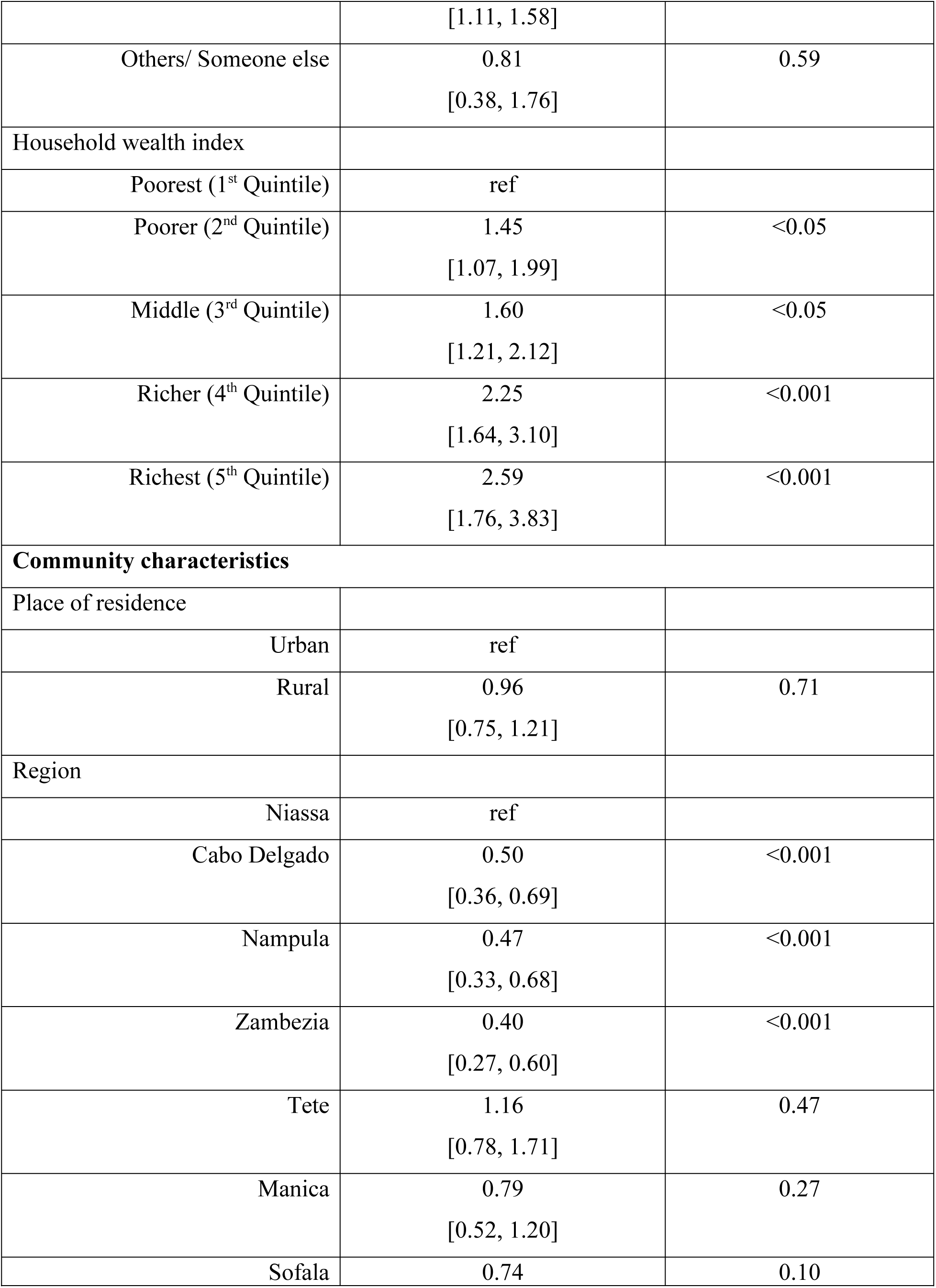
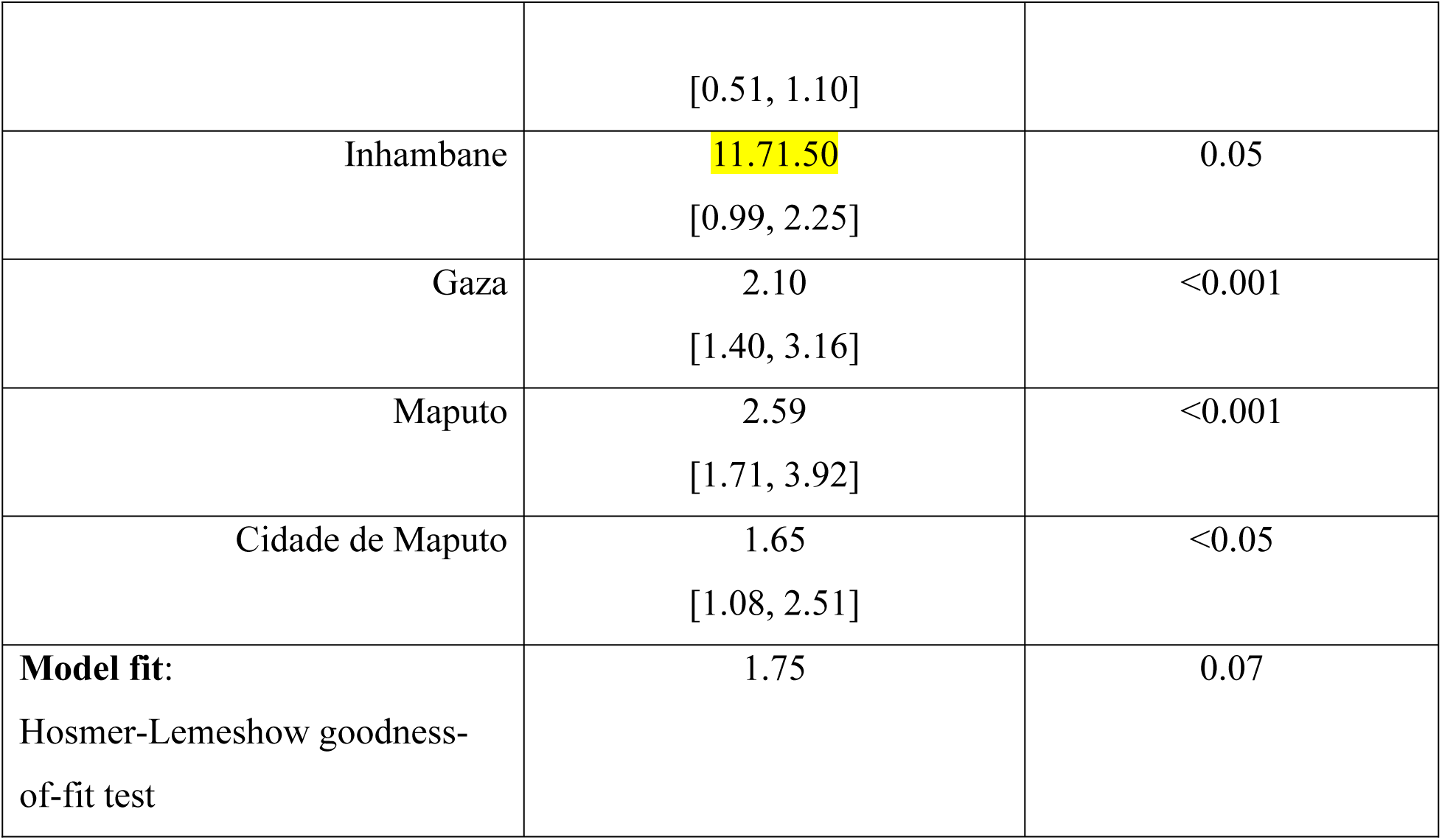
Results from final multivariable binary logistic regression model showing associated factors of modern contraceptive use among reproductive-aged women in Mozambique: DHS2022-23.

Age was significantly associated with contraceptive use. Compared to adolescents aged 15-19 years, women aged 40-44 [aOR = 0.48, 95% CI: 0.34–0.68; p<0.001] and 45–49 [aOR = 0.29, 95% CI: 0.19–0.44; p<0.001] were significantly less likely to use modern contraceptives, indicating a decline in utilization with increasing age. Educational attainment showed a strong positive association. Women with secondary education had over twice the odds of using modern contraceptives [aOR = 2.13, 95% CI: 1.63–2.78; p< 0.001] compared to those with no formal education. Employment status was also significant, with employed women more likely to use contraceptives [aOR = 1.31, 95% CI: 1.11–1.54; p< 0.05]. Religious beliefs influenced contraceptive use. Women who follow Islam had lower odds [aOR = 0.76, 95% CI: 0.59–0.98; p < 0.05] compared to Catholics.

Regarding sexual debut, those initiating sex at 15-19 years had marginally higher odds [aOR= 1.17, 95% CI: 1.00, 1.36; p=0.05] of use compared to those who initiated sex before 15 years of age. Parity was a strong predictor, in a gradient manner. Women with 1-2 children [aOR = 8.94, 95% CI: 5.80, 13.78; p<0.001], 3-4 children [aOR = 14.52, 95% CI: 9.19, 22.96; p<0.001], and ≥5 children [aOR = 23.53, 95% CI: 14.25, 38.84; p<0.001] had significantly higher increasing odds of contraceptive use compared to nulliparous women. Exposure to family planning related information via media was a significant facilitator [aOR = 1.42, 95% CI: 1.19–1.69; p< 0.001]. Joint decision-making with partners also increased the likelihood of use [aOR = 1.32, 95% CI: 1.11-1.58; p< 0.05], compared to decisions made solely by the respondent. Household wealth was a strong determinant. Women in the richest quintile had the highest odds of contraceptive use [aOR = 2.59, 95% CI: 1.76–3.83, p< 0.001] compared to those in the poorest quintile.

Considering community characteristics, place of residence (urban vs. rural) was not significant. However, regional disparities were notable. Women in Cabo Delgado (aOR= 0.50, 95% CI: 0.36, 0.69, p< 0.001), Nampula (aOR= 0.47, 95% CI: 0.33, 0.68, p< 0.001), and Zambezia (aOR= 0.40, 95% CI: 0.27, 0.60, p< 0.001) had significantly lower odds of contraceptive use, while those in Maputo (aOR= 2.59, 95% CI: 1.71, 3.92, p< 0.001) and Gaza (aOR= 2.10, 95% CI: 0.27, 0.60, p< 0.001) had significantly higher odds. The final model demonstrated a good fit to data in this analysis (Hosmer-Lemeshow test: χ² = 1.47, p = 0.15).

### 3.4 Inequalities in utilization of modern contraceptives

**Table 3** represents Erreygers Concentration Index (ECI) values assessing inequality in modern contraceptive use among reproductive-aged women in Mozambique. The ECI is appropriate for binary outcomes and adjusts for the fact that contraceptive use can take only values between 0 and 1, allowing a meaningful interpretation of inequality.

**Table 3:**
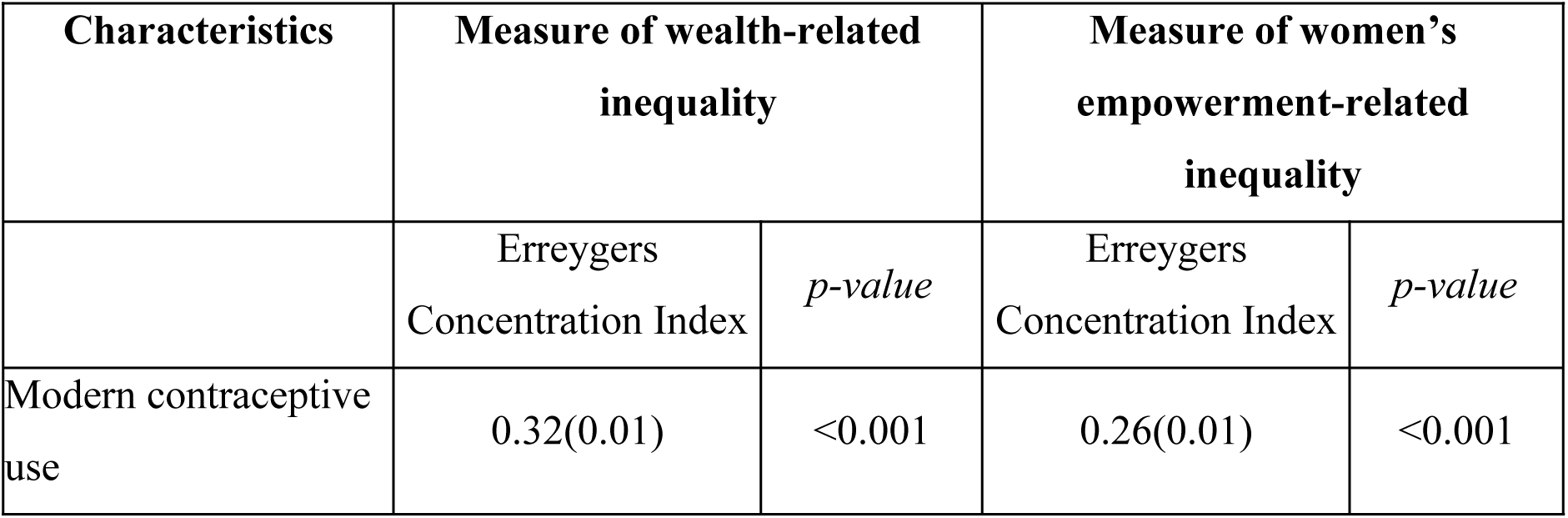
Inequalities related to household wealth and women’s empowerment in modern contraceptive use among reproductive-aged women in Mozambique.

| Characteristics | Measure of wealth-related inequality |  | Measure of women's empowerment-related inequality |  |
| --- | --- | --- | --- | --- |
|  | Erreygers Concentration Index | *p-value* | Erreygers Concentration Index | *p-value* |
| Modern contraceptive use | 0.32(0.01) | <0.001 | 0.26(0.01) | <0.001 |

The wealth-related ECI of 0.32 (SE = 0.01, p< 0.001) indicates a significant pro-rich inequality, meaning women from wealthier households are more likely to use modern contraceptives.

Similarly, the empowerment-related ECI of 0.26 (SE = 0.01, p< 0.001) reflects a pro-empowered distribution, suggesting that women with greater agency are more likely to adopt modern contraceptive methods.

**Figure 2** displays the concentration curves for modern contraceptive use by (a) household wealth and (b) women’s empowerment. Both curves lie below the diagonal line of equality, confirming inequality. The curve for wealth (**Figure 2a**) shows a greater deviation from the diagonal line of equality, consistent with the higher ECI value and indicating higher household wealth inequality of modern contraception use. These findings, however, highlight the dual influence of wealth status and women’s empowerment on modern contraceptive uptake, underscoring the need for equity-oriented reproductive health strategies.

**Figure 2:**
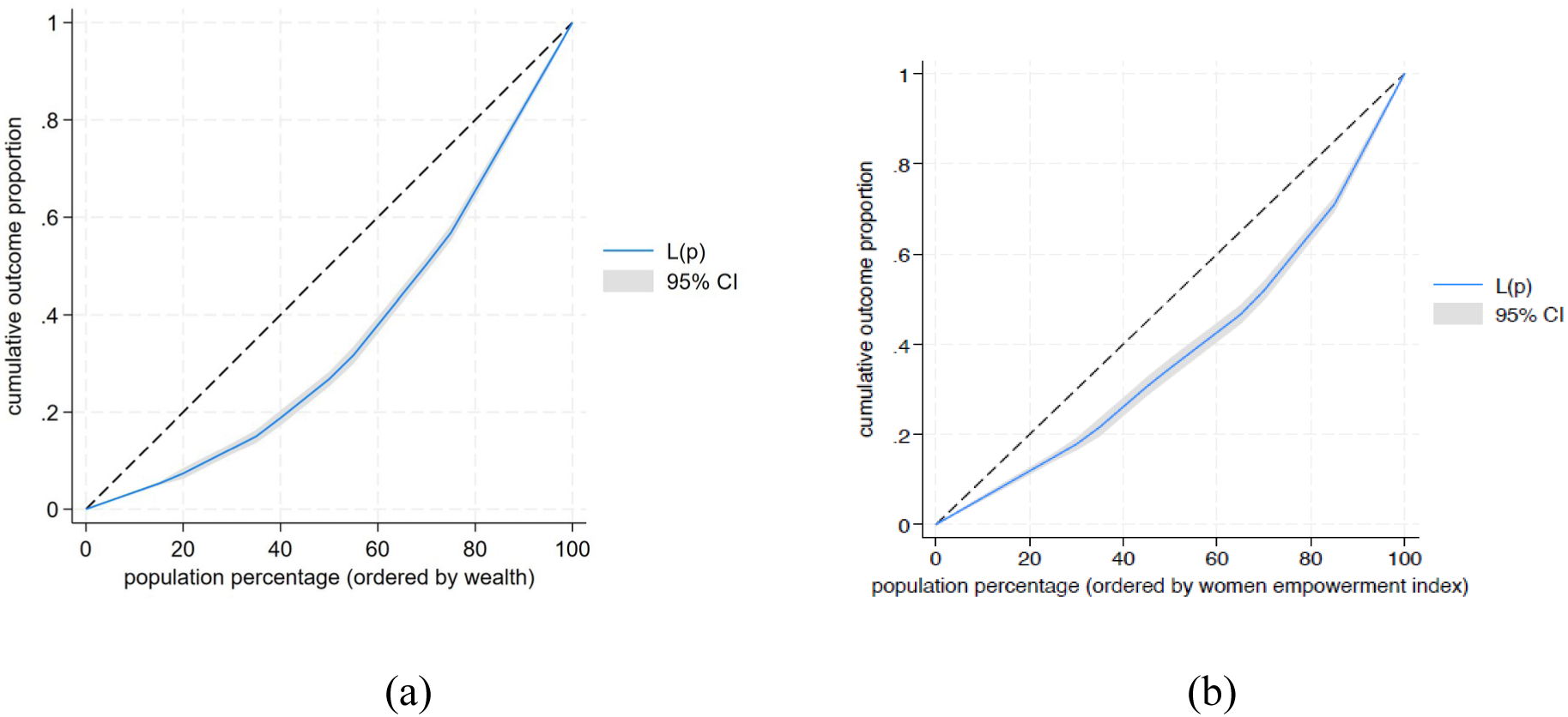
Concentration curves of modern contraceptive use based on (a) household wealth status and (b) women’s empowerment. Note: L(p) refers to cumulative proportion.

## DISCUSSION

Our study examined the prevalence of modern contraceptive use among women of reproductive age (15-49 years) in Mozambique. The findings underscore the interplay between sociodemographic, household-related, and community factors influencing women’s contraceptive choices. Overall, 27.6% of the study participants reported use of modern contraceptives. This prevalence is comparable to the findings from previous studies conducted in Ethiopia,(27) Kenya,(28) and Uganda,(29) suggesting that modern contraceptive uptake in Mozambique aligns with patterns observed across similar sub-Saharan African settings.

Our study found an inverse association between age and modern contraceptive use, indicating that younger women were more likely to use contraceptives compared to older women. Similar age-related trends have been documented in other sub-Saharan African settings,(30–33) where contraceptive uptake declines as women approach the end of their reproductive life. This tendency may reflect decreasing perceived need for contraception due to decreasing reproductive intentions as women age.(34) Additionally, awareness of age-related declines in fecundity, or a sense of complacency about pregnancy risk may contribute to reduced contraceptive use among older women.

In our study, education was identified as a strong positive determinant of modern contraceptive use, consistent with findings from previous studies in Mozambique and other LMICs. (30,33,35,36) Women with higher levels of education are more likely to have greater knowledge of contraceptive methods, reproductive health, and access to health services, which enhances their capacity to make informed reproductive decisions.(37,38) Education may also empower women to negotiate contraceptive use within relationships and challenge traditional gender norms that restrict women’s autonomy, which facilitates higher utilization of modern contraceptives.

Furthermore, being employed was associated with higher contraceptive use, suggesting that economic empowerment contributes to greater agency and decision-making in reproductive matters. Gebre et al.(39) found that women engaging in various economic activities like selling goods, agricultural work, skilled manual labor, and other jobs were more likely to use modern contraceptives in Ethiopia. These findings may reflect improved access to information, mobility, and health services, as well as a stronger desire to regulate fertility to align with career or economic aspirations.(40,41)

Household wealth was another strong determinant, with women in the richest households having more than twice the odds of using modern contraceptives compared to those in the poorest. Similar findings were reported by Tesema et al.(42) in their analysis of DHS data from 36 sub-Saharan African countries. Women’s capacity to purchase modern contraceptives, themselves, and not necessarily relying on their partners is the likely reason for this finding.

Our study found lower odds of modern contraceptive use among women who professes Islam compared to Catholics, which may reflect religious and cultural influences on reproductive preferences and attitude toward contraception. Religion has been a strong predictor of contraceptive use in many studies.(36,41,43,44) In some contexts, Islamic teachings are interpreted as discouraging modern contraceptive methods, particularly those viewed as interfering with natural fertility. Community-level religious norms and social expectations may further reinforce these beliefs, contributing to reduced uptake of modern contraceptives among Muslim women.

Our findings indicate that women’s parity has a pivotal role in shaping modern contraceptive use. The progressively higher odds with increasing number of children in our study indicate that women are more likely to adopt contraceptive use after achieving desired family size, aligning with birth spacing to limiting fertility transition.(45–47) This finding highlights the need for contraceptive programs to also target nulliparous and low-parity women emphasizing the health and socioeconomic benefits of birth spacing rather than limiting births.

Partner’s education also played a significant role in utilizing modern contraceptives in Mozambique. Similarly, Gebre and Edossa(48) reported that women whose husbands had primary education were 40% more likely to use modern contraceptives compared to those whose husbands had no formal education. This may be because educated partners are better informed about modern contraception, less influenced by restrictive social norms, and more supportive of their wives’ reproductive choices.

Exposure to family planning information through media was a significant factor for contraceptive use, underscoring the importance of mass media campaigns in promoting awareness and shaping attitudes. Media exposure has been widely recognized as an effective avenue for disseminating family planning messages, countering misinformation, and normalizing contraceptive use.(30,36,49) Likewise, joint decision-making with partners significantly increased contraceptive uptake, reinforcing the role of male involvement and couple communication in reproductive health behaviors.(50) Programs that encourage spousal communication and shared decision-making may therefore be effective in increasing uptake.

Contrary to expectations, place of residence (urban vs. rural) was not significantly associated with modern contraceptive use after adjusting for other factors. Nevertheless, regional disparities are evident, with provinces such as Cabo Delgado, Nampula, and Zambezia exhibiting significantly lower contraceptive use. These provinces have historically faced challenges related to limited health infrastructure, social and cultural barriers, and political insecurity,(51) which may restrict access to and acceptance of family planning services. In contrast, higher odds in Maputo and Gaza likely reflect better service availability, stronger local health systems, and higher literacy rates.

Our study highlights the significant wealth-related and empowerment-related inequalities in modern contraceptive use among women in Mozambique. The wealth-related ECI of 0.32 indicates that women from wealthier households are more likely to use modern contraceptives, while the empowerment-related ECI of 0.26 shows higher uptake among women with greater empowerment level. These results align with studies from other LMICs, highlighting the roles of both economic resources and women’s autonomy in shaping contraceptive behavior.(52–54) These findings underscore the determinant power of two core contextual determinants, household wealth and women’s empowerment, and the need for equity-focused interventions that address both financial barriers and women’s empowerment to promote inclusive access to modern family planning services.

### Strength and weaknesses

The strengths include the use of nationally representative data, a large sample size providing robust estimates of modern contraceptive use and its associated factors. This analysis also incorporated sample weighting, which enhances the statistical power and generalizability. Given that the DHS employs a standardized design with identical variables across different contexts, the findings may be applicable to other similar locations. However, this study has several limitations. The cross-sectional design limits the ability to draw causal inferences. Data are based on self-reported measures, which may be subject to recall bias. However, inclusion of questions focused on current contraceptive use at the time of the survey mitigates this concern.

Additionally, cultural norms and social behaviors related to women’s reproductive decision making and contraceptive utilization could not be fully captured due to lack of relevant variables in the DHS data.

## CONCLUSION

Our study analyzed the Mozambvique DHS 2022-2023 data, focusing on modern contraceptive utilization among women of reproductive age. Our findings highlights that modern contraceptive use among women of reproductive age in Mozambique remains suboptimal and is associated with a range of sociodemographic, reproductive, and contextual factors, including age, formal education, employment, parity, wealth status, and partner characteristics. Significant inequalities were observed, with wealthier and more empowered women disproportionately benefiting from accessing modern contraceptive methods. These findings underscore the need for multifaceted, equitable interventions that can address financial, informational, and sociocultural barriers.

Achieving universal access to sexual and reproductive health services, as emphasized in the 2030 Agenda for Sustainable Development, requires coordinated efforts from governments, NGOs, and other stakeholders to strengthen family planning programs in Mozambique. Aligning national efforts with global initiatives, such as WHO guidelines and support for contraceptive innovation, can further improve access, utilization, and the overall effectiveness of reproductive health services. Policy efforts should prioritize expanding access to family planning services for economically disadvantaged and low-empowerment women, promoting women’s autonomy in reproductive decision-making, and integrating media and community-based education campaigns to improve awareness. Strengthening spousal communication and male partners involvement, alongside targeting low-parity and younger women, can further improve contraceptive uptake. By addressing both individual and structural determinants, policymakers can enhance reproductive health equity and progress toward national and global family planning targets.

## Data Availability

All data produced in the present study are available upon reasonable request to the authors.

## Acknowledgement

We greatly acknowledge the Demographic and Health Survey (DHS) program for granting access to the Mozambique DHS datasets.

## DECLARATIONS

### Author contributions

NM and NS conceptualized this study. NS performed formal analysis of the data. NS (methodology, results, discussion) and FA (background) wrote the first draft. All authors reviewed, edited the drafts and approved the final version of the manuscript. NM supervised the work and holds provenance for the article.

### Ethics declaration

This study used data from Mozambique Demographic and Health Survey 2022-2023. The survey was approved by National Bioethics Committee for Health (CNBS) and ICF International Institutional Review Board (IRB). Further approval for this study was not required since data is freely available in the public domain.

### Data availability statement

The Mozambique Demographic and Health Survey data is available from the DHS data repository which is publicly available.

### Patient and public involvement

Patients and/or the public were not involved in the design, or conduct, or reporting, or dissemination plans of our study.

## REFERENCES

1. Cook RJ, Dickens BM, Fathalla MF. Reproductive Health and Human Rights: Integrating Medicine, Ethics, and Law [Internet]. Oxford University Press; 2003 [cited 2025 Oct 6]. Available from: 10.1093/acprof:oso/9780199241323.001.0001

2. Black RE, Walker N, Laxminarayan R, Temmerman M. Reproductive, Maternal, Newborn, and Child Health: Key Messages of This Volume. In: Black RE, Laxminarayan R, Temmerman M, Walker N, editors. Reproductive, Maternal, Newborn, and Child Health: Disease Control Priorities, Third Edition (Volume 2) [Internet]. Washington (DC): The International Bank for Reconstruction and Development / The World Bank; 2016 [cited 2025 Oct 6]. Available from: http://www.ncbi.nlm.nih.gov/books/NBK361926/

3. Anaba EA, Wright ME, Alor SK, Lindeman M, Salifu Y, Adjorlolo S, et al. Contraceptive use and mental health among women of reproductive age: insights from the Mozambique Demographic and Health Survey. BJPsych Int. 2025 July 11;1–7.

4. United Nations, Department of Economic and Social Affairs, Population Division (2019). Contraceptive Use by Method 2019: Data Booklet (ST/ESA/SER.A/435). [Internet]. 2019. Available from: http://un-ilibrary.org/content/books/9789210046527

5. Cardona C, Rusatira JC, Salmeron C, Martinez-Baack M, Rimon JG, Anglewicz P, et al. Progress in reducing socioeconomic inequalities in the use of modern contraceptives in 48 focus countries as part of the FP2030 initiative between 1990 and 2020: a population-based analysis. Lancet Glob Health. 2025 Jan 1;13(1):e38–49.

6. Ntoimo L. Contraception in Africa: Is the global 2030 milestone attainable? Afr J Reprod Health [Internet]. 2021 June 28 [cited 2025 Oct 6];25(3). Available from: https://www.ajrh.info/index.php/ajrh/article/view/2772

7. Group BMJP. Maternal Morbidity and Mortality: A Scottish Report. Br Med J. 1935 Aug 10;2(3892):265–7.

8. Dias JG, Oliveira IT de. Multilevel Effects of Wealth on Women’s Contraceptive Use in Mozambique. PLOS ONE. 2015 Mar 18;10(3):e0121758.

9. Amoah EJ, Hinneh T, Aklie R. Determinants and prevalence of modern contraceptive use among sexually active female youth in the Berekum East Municipality, Ghana. PLOS ONE. 2023 June 8;18(6):e0286585.

10. Ahinkorah BO, Budu E, Aboagye RG, Agbaglo E, Arthur-Holmes F, Adu C, et al. Factors associated with modern contraceptive use among women with no fertility intention in sub-Saharan Africa: evidence from cross-sectional surveys of 29 countries. Contracept Reprod Med. 2021 Aug 1;6(1):22.

11. Family planning/contraception methods [Internet]. [cited 2025 Oct 6]. Available from: https://www.who.int/news-room/fact-sheets/detail/family-planning-contraception

12. Daca C, Sebastian MS, Arnaldo C, Schumann B. Socio-economic and demographic factors associated with reproductive and child health preventive care in Mozambique: a cross-sectional study. Int J Equity Health. 2020 Nov 9;19(1):200.

13. The DHS Program - Mozambique: Standard DHS, 2011 [Internet]. [cited 2025 Oct 6]. Available from: https://dhsprogram.com/methodology/survey/survey-display-362.cfm

14. Misau M da S, Ine IN de E, ICF. Inquérito de Indicadores de Imunização, Malária e HIV/SIDA em Moçambique (IMASIDA) 2015. 2018 Feb 1 [cited 2025 Oct 6]; Available from: https://dhsprogram.com/publications/publication-ais12-ais-final-reports.cfm

15. Contraception: the key to achieving SDGs by 2030 | Figo [Internet]. 2018 [cited 2025 Oct 6]. Available from: https://www.figo.org/contraception-key-achieving-sdgs-2030

16. SDG Indicator 3.7.1 on Contraceptive Use | Population Division [Internet]. [cited 2025 Oct 6]. Available from: https://www.un.org/development/desa/pd/data/sdg-indicator-371-contraceptive-use

17. Wulifan JK, Mazalale J, Kambala C, Angko W, Asante J, Kpinpuo S, et al. Prevalence and determinants of unmet need for family planning among married women in Ghana-a multinomial logistic regression analysis of the GDHS, 2014. Contracept Reprod Med. 2019 Jan 30;4(1):2.

18. Gueye A, Speizer IS, Corroon M, Okigbo CC. Belief in Family Planning Myths at the Individual And Community Levels and Modern Contraceptive Use in Urban Africa. Int Perspect Sex Reprod Health. 2015 Dec;41(4):191–9.

19. Belda SS, Haile MT, Melku AT, Tololu AK. Modern contraceptive utilization and associated factors among married pastoralist women in Bale eco-region, Bale Zone, South East Ethiopia. BMC Health Serv Res. 2017 Mar 14;17(1):194.

20. National Institute of Statistics (INE) and ICF. 2024. Mozambique Demographic and Health survey 2022-23 [Internet]. The DHS Program ICF Rockville, Maryland, USA; 2024 May. Available from: https://dhsprogram.com/publications/publication-FR389-DHS-Final-Reports.cfm

21. The DHS Program - Mozambique: Standard DHS, 2022-23 [Internet]. [cited 2024 Sept 18]. Available from: https://dhsprogram.com/methodology/survey/survey-display-564.cfm

22. Hubacher D, Trussell J. A definition of modern contraceptive methods. Contraception. 2015 Nov;92(5):420–1.

23. Ewerling F, Raj A, Victora CG, Hellwig F, Coll CV, Barros AJ. SWPER Global: A survey-based women’s empowerment index expanded from Africa to all low-and middle-income countries. J Glob Health. 10(2):020434.

24. Croft TN, Allen CK, Zachary BW, et al. Guide to DHS Statistics [Internet]. Rockville, Maryland, USA: ICF; 2023. Available from: https://dhsprogram.com/data/Guide-to-DHS-Statistics/index.cfm

25. UNU-WIDER : Book : Health Inequality and Development [Internet]. [cited 2024 Aug 8]. Available from: http://www.wider.unu.edu/publication/health-inequality-and-development-0

26. Erreygers G. Correcting the Concentration Index. J Health Econ. 2009 Mar 1;28(2):504–15.

27. Merera AM, Lelisho ME, Sheferaw WE. Determinants of modern contraceptive use among married women of reproductive age in ethiopia: a cross-sectional analysis of the 2019 Ethiopian mini demographic and health survey. Sci Rep. 2025 Oct 9;15(1):35290.

28. Lunani LL, Abaasa A, Omosa-Manyonyi G. PREVALENCE AND FACTORS ASSOCIATED WITH CONTRACEPTIVE USE AMONG KENYAN WOMEN AGED 15– 49 YEARS. AIDS Behav. 2018 July;22(Suppl 1):125–30.

29. Asiimwe JB, Ndugga P, Mushomi J, Manyenye Ntozi JP. Factors associated with modern contraceptive use among young and older women in Uganda; a comparative analysis. BMC Public Health. 2014 Sept 8;14(1):926.

30. Boadu I. Coverage and determinants of modern contraceptive use in sub-Saharan Africa: further analysis of demographic and health surveys. Reprod Health. 2022 Jan 21;19(1):18.

31. Zeleke GT, Zemedu TG. Modern contraception utilization and associated factors among all women aged 15–49 in Ethiopia: evidence from the 2019 Ethiopian Mini Demographic and Health Survey. BMC Womens Health. 2023 Feb 9;23(1):51.

32. Forty J, Rakgoasi SD, Keetile M. Patterns and determinants of modern contraceptive use and intention to usecontraceptives among Malawian women of reproductive ages (15–49 years). Contracept Reprod Med. 2021 July 1;6:21.

33. Lahole BK, Banga D, Mare KU. Modern contraceptive utilization among women of reproductive age in Ghana: a multilevel mixed-effect logistic regression model. Contracept Reprod Med. 2024 Sept 27;9(1):46.

34. Bandehelahi K, Khoshravesh S, Barati M, Tapak L. Psychological and Sociodemographic Predictors of Fertility Intention among Childbearing-Aged Women in Hamadan, West of Iran: An Application of the BASNEF Model. Korean J Fam Med. 2019 May;40(3):182–7.

35. Mutumba M, Wekesa E, Stephenson R. Community influences on modern contraceptive use among young women in low and middle-income countries: a cross-sectional multi-country analysis. BMC Public Health. 2018 Apr 2;18(1):430.

36. Demeke H, Legese N, Nigussie S. Modern contraceptive utilization and its associated factors in East Africa: Findings from multi-country demographic and health surveys. PLOS ONE. 2024 Jan 19;19(1):e0297018.

37. Hahn RA, Truman BI. Education Improves Public Health and Promotes Health Equity. Int J Health Serv Plan Adm Eval. 2015;45(4):657–78.

38. Gelgelo D, Abeya SG, Hailu D, Edin A, Gelchu S. Effectiveness of Health Education Interventions Methods to Improve Contraceptive Knowledge, Attitude, and Uptake Among Women of Reproductive Age, Ethiopia: A Systematic Review and Meta-Analysis. Health Serv Res Manag Epidemiol. 2023 Jan 1;10:23333928221149264.

39. Gebre G, Birhan N, Gebreslasie K. Prevalence and factors associated with unmet need for family planning among the currently married reproductive age women in Shire-Enda-Slassie, Northern West of Tigray, Ethiopia 2015: a community based cross-sectional study. Pan Afr Med J [Internet]. 2016 July 14 [cited 2024 June 5];23(1). Available from: https://www.ajol.info/index.php/pamj/article/view/139530

40. Tekelab T, Melka AS, Wirtu D. Predictors of modern contraceptive methods use among married women of reproductive age groups in Western Ethiopia: a community based cross-sectional study. BMC Womens Health. 2015 July 17;15(1):52.

41. Adebowale AS, Gbadebo B, Afolabi FR. Wealth index, empowerment and modern contraceptive use among married women in Nigeria: are they interrelated? J Public Health. 2016 Oct 1;24(5):415–26.

42. Tesema ZT, Tesema GA, Boke MM, Akalu TY. Determinants of modern contraceptive utilization among married women in sub-Saharan Africa: multilevel analysis using recent demographic and health survey. BMC Womens Health. 2022 May 18;22(1):181.

43. Tiruneh FN, Chuang KY, Ntenda PAM, Chuang YC. Factors associated with contraceptive use and intention to use contraceptives among married women in Ethiopia. Women Health. 2016;56(1):1–22.

44. Lasong J, Zhang Y, Gebremedhin SA, Opoku S, Abaidoo CS, Mkandawire T, et al. Determinants of modern contraceptive use among married women of reproductive age: a cross-sectional study in rural Zambia. BMJ Open. 2020 Mar 1;10(3):e030980.

45. Bongaarts J, Casterline J. Fertility Transition: Is sub-Saharan Africa Different? Popul Dev Rev. 2013 Feb;38(Suppl 1):153–68.

46. Mostafa Kamal SM, Aynul Islam M. Contraceptive Use: Socioeconomic Correlates and Method Choices in Rural Bangladesh. Asia Pac J Public Health. 2010 Oct 1;22(4):436–50.

47. Zegeye B, Ahinkorah BO, Idriss-Wheeler D, Olorunsaiye CZ, Adjei NK, Yaya S. Modern contraceptive utilization and its associated factors among married women in Senegal: a multilevel analysis. BMC Public Health. 2021 Jan 28;21(1):231.

48. Gebre MN, Edossa ZK. Modern contraceptive utilization and associated factors among reproductive-age women in Ethiopia: evidence from 2016 Ethiopia demographic and health survey. BMC Womens Health. 2020 Mar 26;20(1):61.

49. Sserwanja Q, Turimumahoro P, Nuwabaine L, Kamara K, Musaba MW. Association between exposure to family planning messages on different mass media channels and the utilization of modern contraceptives among young women in Sierra Leone: insights from the 2019 Sierra Leone Demographic Health Survey. BMC Womens Health. 2022 Sept 16;22(1):376.

50. Do M, Kurimoto N. Women’s Empowerment and Choice of Contraceptive Methods in Selected African Countries. Int Perspect Sex Reprod Health. 2012 Mar 30;38:23.

51. Muhajarine N, Shakurun N, Ahmed MS, Andre F, Chicumbe S. Inequalities and factors associated with maternal healthcare services utilisation in Mozambique: evidence from the Demographic and Health Survey 2022−2023. BMJ Glob Health [Internet]. 2025 May 24 [cited 2025 Aug 25];10(5). Available from: https://gh.bmj.com/content/10/5/e018121

52. Kazibwe J, Masiye F, Klingberg-Allvin M, Ekman B, Sundewall J. Inequality in modern contraceptive use and unmet need for contraception among women of reproductive age in Zambia. A trend and decomposition analysis 2007–2018. Reprod Health. 2024 Dec 9;21(1):181.

53. Budu E, Dadzie LK, Salihu T, Ahinkorah BO, Ameyaw EK, Aboagye RG, et al. Socioeconomic inequalities in modern contraceptive use among women in Benin: a decomposition analysis. BMC Womens Health. 2023 Aug 23;23(1):444.

54. Srivastava S, Mohanty P, Muhammad T, Kumar M. Socio-economic inequalities in non-use of modern contraceptives among young and non-young married women in India. BMC Public Health. 2023 May 1;23(1):797.

