## Supplementary material for "Predictors of modern contraceptive use among reproductive-aged women (15-49 years) in Mozambique: Evidence from Mozambique Demographic and Health Survey"

### Supplementary file

These supplementary materials are in support of the paper:
Predictors of modern contraceptive use among reproductive aged women (15-49 years) in Mozambique: Evidence from Mozambique Demographic and Health Survey.
Nahin Shakurun, Fernanda Andre, Nazeem Muhajarine

**Sample selection:**

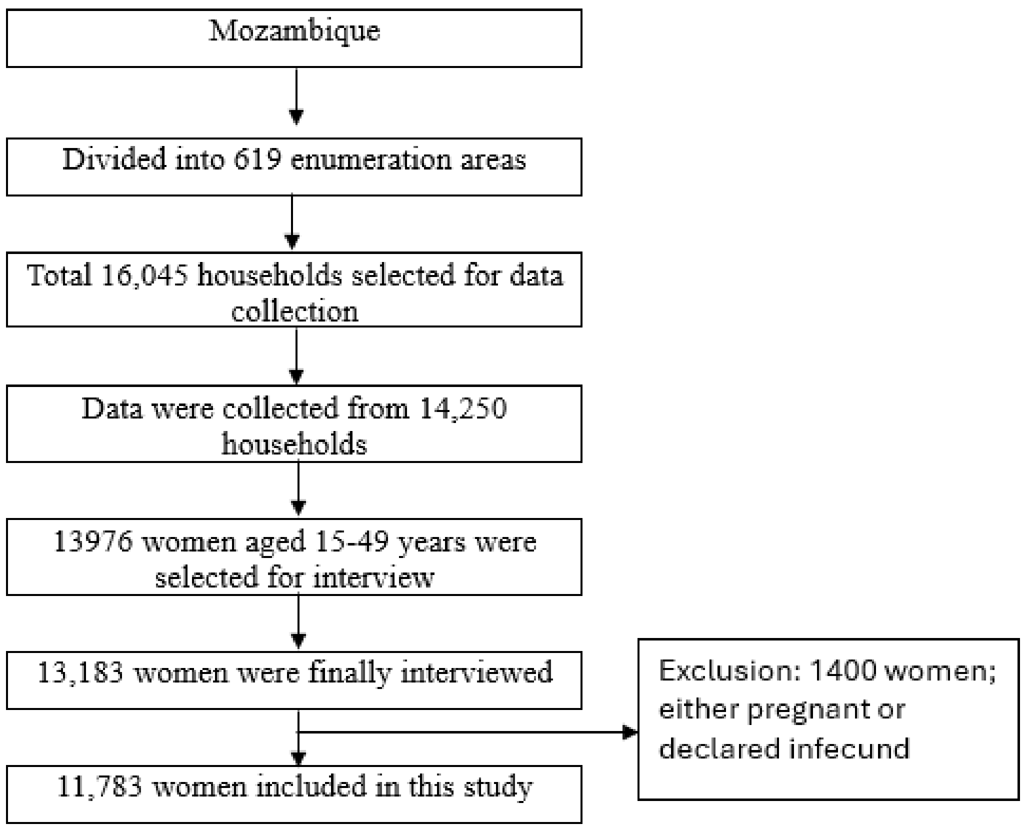

**Figure S1:** Selection of participants for the study of modern contraceptive use by reproductive age women in Mozambique.

**Table S1:** Independent variables and their categorization

| Variable | Description | Categorization |
|---|---|---|
| **Individual characteristics** | | |
| Age | Age of the respondents(women) in years | 0. 15-19 years<br>1. 20-24 years |

| | | |
|---|---|---|
| | | 2. 25-29 years<br>3. 30-34 years<br>4. 35-39 years<br>5. 40-44 years<br>6.45-49 years |
| Marital/ Partnered status | Current marital status of the respondents | 0. Never in union<br>1. Partnered (married/living with partner)<br>2. Non-partnered (widowed, divorced, separated) |
| Education | Highest educational grades completed by the respondent | 0. No formal education<br>1. Primary<br>2. Secondary<br>3. Higher |
| Employment status | Current working status of the respondents | 0. Not working<br>1. Working |
| Religion | Religion | 0. Catholic<br>1. Islamic<br>2. Zion<br>3. Evangelical/Pentecostal<br>4. Others |
| Women empowerment index | Women empowerment index was developed using principal component analysis. Details about this index are provided in Table 2. | 0. Lowest<br>1. Low<br>2. Moderate<br>3. High<br>4. Highest |
| Age at first sex | Age at first sex | 0. <15 years<br>1. 15-19 years<br>2. 20 years and above |
| No. of children born alive | Number of children who are currently alive | 0. No children |

| | | |
|---|---|---|
| | | 1. 1-2<br>2. 3-4<br>3. 5 or more |
| Husband's education | Husband/partner's highest educational grades of the respective respondent. | 0. No formal education<br>1. Primary<br>2. Secondary<br>3. Higher |
| Decision maker for the contraceptive use | Decision maker for the contraceptive use | 0. Respondent<br>1. Husband/partner<br>2. Joint decision<br>3. Others |
| Household wealth index | Household wealth status | 0. Poorest<br>1. Poorer<br>2. Middle<br>3. Richer<br>4. Richest |
| Heard family planning on media | Heard about family planning on media like radio, TV, newspaper or magazine in last few months | 0. No<br>1. Yes |
| **Community characteristics** | | |
| Place of residence | Type of place of residence | 0. Urban<br>1. Rural |
| Region | Administrative regions | 0. Maputo<br>1. Niassa<br>2. Cabo Delgado<br>3. Nampula<br>4. Zambezia<br>5. Tete<br>6. Manica<br>7. Sofala |

| | | |
|---|---|---|
| | | 8. Inhambane<br>9. Gaza<br>11. Cidade de Maputo |

**Table S2:** Variables considered to create women empowerment index: Mozambique Demographic and Health Survey Data 2022-23

| Variable | Categorization | Re-categorization |
|---|---|---|
| 1. Person who usually decides on: respondent's health care | 1. Alone<br>2. Jointly with husband/partner<br>3. Jointly with other person<br>4. Husband alone<br>5. Someone else<br>6. other | • Alone and jointly with husband/partner= 1<br>• Jointly with other person, husband alone, someone else, other= 0 |
| 2. Person who usually decides on: large household purchases | | |
| 3. Person who usually decides on: visits to family or relatives | | |
| 4. Beating justified if wife goes out without telling husband | 0. No<br>1. Yes<br>8. Don't know | • No= 1<br>• Yes and don't know= 0 |
| 5. Beating justified if wife neglects the children | | |
| 6. Beating justified if wife argues with husband | | |
| 7. Beating justified if wife refuses to have sex with husband | | |
| 8. Beating justified if wife burns the food | | |
| 9. Respondent currently working | 0. No<br>1. Yes | • No= 0<br>• Yes= 1 |
| 10. Has an account in a bank or other financial institution | | |
| 11. Owns a mobile/telephone | | |
| 12. Reading newspaper or Magazine at least once | | |

| | | |
|---|---|---|
| a week | | |
| 13. Owns a house/land alone or jointly | 0. Does not own<br>1. Alone only<br>2. Jointly with husband/partner<br>3. Jointly with someone else<br>4. Jointly with husband/partner and someone else<br>5. Both alone and jointly | • Does not own= 0<br>• Own= 1 |

**Table S3:** Prevalence of modern method of contraceptive use by reproductive age women in Mozambique

| Characteristics | N (Unweighted) | % (Weighted) |
|---|---|---|
| Modern contraceptives | | |
| No modern method used | 7923 | 72.4 |
| Modern method used | 3860 | 27.6 |

**Table S4:** Bivariable analysis of modern contraceptive use by reproductive age women in Mozambique

| Characteristics | Utilization of modern contraceptive | |
|---|---|---|
| | Yes<br>% [95% CI] | p-value |
| **Individual characteristics** | | |
| Age | | <0.001 |
| 15-19 years | 17.21<br>[15.58, 18.97] | |
| 20-24 years | 31.36<br>[28.78, 34.07] | |
| 25-29 years | 31.85<br>[29.01, 34.83] | |

| | | |
|---|---|---|
| 30-34 years | 35.59 [32.49, 38.81] | |
| 35-39 years | 34.12 [30.76, 37.64] | |
| 40-44 years | 28.11 [25.02, 31.42] | |
| 45-49 years | 17.17 [14.67, 20.00] | |
| Education status | | <0.001 |
| No formal education | 12.51 [10.93, 14.28] | |
| Primary | 24.50 [22.76, 26.33] | |
| Secondary | 43.41 [41.13, 45.71] | |
| Post-secondary | 49.88 [44.76, 55.00] | |
| Employment status | | <0.001 |
| Unemployed | 21.92 [20.39, 23.54] | |
| Employed | 40.45 [38.22, 42.72] | |
| Marital status | | <0.001 |
| Never in union | 23.75 [21.80, 25.81] | |
| Partnered women | 28.72 [26.92, 30.59] | |
| Non-partnered (widowed, divorced, separated) | 29.15 [26.59, 31.84] | |
| Religion | | <0.001 |

| | | |
|---|---|---|
| Catholic | 21.36 [19.26, 23.62] | |
| Islamic | 16.22 [13.76, 19.03] | |
| Zion | 34.40 [31.40, 37.54] | |
| Evangelical/Pentecostal | 38.81 [36.40, 41.27] | |
| Others | 28.79 [25.12, 32.77] | |
| Age at first sex | | <0.001 |
| <15 years | 22.13 [20.61, 23.72] | |
| 15-19 years | 35.04 [33.10, 37.95] | |
| 20 years and above | 28.07 [19.94, 37.95] | |
| No. of living children | | <0.001 |
| No children | 18.26 [16.60, 20.04] | |
| 1-2 children | 31.89 [29.87, 33.94] | |
| 3-4 children | 31.88 [29.35, 34.52] | |
| 5 or more children | 26.40 [23.58, 29.42] | |
| Women empowerment index | | <0.001 |
| Lowest (1st Quintile) | 17.06 [14.81, 19.58] | |
| Low (2nd Quintile) | 25.70 | |

| | | |
|---|---|---|
| | [22.46, 29.24] | |
| Moderate (3rd Quintile) | 22.75<br>[19.95, 25.81] | |
| High (4th Quintile) | 36.52<br>[33.31, 39.86] | |
| Highest (5th Quintile) | 55.71<br>[52.44, 58.92] | |
| Husband's education | | <0.001 |
| No formal education | 14.80<br>[12.80, 17.05] | |
| Primary | 26.75<br>[24.40, 29.24] | |
| Secondary | 47.04<br>[43.67, 50.44] | |
| Higher | 58.24<br>[52.27, 63.97] | |
| Heard family planning on media | | <0.001 |
| No | 18.26<br>[16.89, 19.71] | |
| Yes | 40.99<br>[38.99, 43.02] | |
| Decision maker for contraceptive use | | <0.001 |
| Respondent | 34.09<br>[31.35, 36.95] | |
| Husband/partner | 18.60<br>[16.28, 21.16] | |
| Joint decision | 33.52<br>[30.83, 36.32] | |
| Others/ Someone else | 11.30 | |

| | | |
|---|---|---|
| | [5.94, 20.44] | |
| Household wealth index | | <0.001 |
| Poorest (1st Quintile) | 9.72<br>[8.04, 11.70] | |
| Poorer (2nd Quintile) | 14.10<br>[11.90, 16.63] | |
| Middle (3rd Quintile) | 22.08<br>[19.58, 24.80] | |
| Richer (4th Quintile) | 34.21<br>[31.35, 37.20] | |
| Richest (5th Quintile) | 47.73<br>[45.41, 50.06] | |
| **Community characteristics** | | |
| Place of residence | | <0.001 |
| Urban | 39.19<br>[36.87, 41.56] | |
| Rural | 19.92<br>[18.41, 21.52] | |
| Region | | <0.001 |
| Maputo | 55.04<br>[50.62, 59.38] | |
| Niassa | 23.83<br>[19.54, 28.71] | |
| Cabo Delgado | 16.87<br>[14.58, 19.44] | |
| Nampula | 13.47<br>[10.95, 16.47] | |
| Zambezia | 12.23<br>[9.36, 15.83] | |
| Tete | 29.78 | |

| | |
|---|---|
| | [25.90, 33.96] |
| Manica | 26.20<br>[22.13, 30.73] |
| Sofala | 32.58<br>[28.39, 37.07] |
| Inhambane | 43.79<br>[39.65, 48.02] |
| Gaza | 48.98<br>[45.26, 52.71] |
| Cidade de Maputo | 54.12<br>[50.51, 57.68] |
